# Serological testing reveals the hidden COVID-19 burden among healthcare workers experiencing a SARS-CoV-2 nosocomial outbreak

**DOI:** 10.1101/2021.07.15.21260585

**Authors:** Yu Nakagama, Yuko Komase, Katherine Candray, Sachie Nakagama, Fumiaki Sano, Tomoya Tsuchida, Hiroyuki Kunishima, Takumi Imai, Ayumi Shintani, Yuko Nitahara, Natsuko Kaku, Yasutoshi Kido

## Abstract

We describe the results of testing healthcare workers from a tertiary care hospital in Japan, which had experienced a COVID-19 outbreak during the first peak of the pandemic, for SARS-CoV-2 specific antibody seroconversion. Using two chemiluminescent immunoassays and a confirmatory surrogate virus neutralization test, serological testing unveiled that a surprising 42.2% (27/64) of overlooked COVID-19 diagnoses had occurred when case detection had relied solely on SARS-CoV-2 nucleic acid amplification testing. This undetected portion of the COVID-19 iceberg beneath the surface may potentially have led to silent transmissions and triggered the spread. A questionnaire-based risk assessment was further indicative of exposures to specific aerosol-generating procedures, i.e. non-invasive ventilation, having had conveyed the highest transmission risks and served as the origin of outbreak. Our observations are supportive of a multi-tiered testing approach, including the use of serological diagnostics, in order to accomplish exhaustive case detection along the whole COVID-19 spectrum.

## Introduction

When the COVID-19 pandemic landed in January 2020, Japan was no exception to the rest of the world, where access to diagnostic testing was limited. Shortages in testing resources during the first wave of the pandemic in spring 2020 had compromised timely case detection and forced healthcare workers (HCWs) to work in a deep diagnostic fog. The situation caused frontline healthcare facilities to suffer unexpected SARS-CoV-2 exposures followed by nosocomial outbreaks. However, even after a profound increase in molecular testing capacity and an apparent clearance of the fog, the SARS-CoV-2 virus continued to sneak through the shield of symptom-driven screening strategies (*1*). Infections free of symptoms, i.e. pre-symptomatic or asymptomatic infections, and thus left untested were hypothesized to constitute a major burden and contribute to transmission (*2*).

In support of the hypothesis, reports from later active screening studies have revealed a significant majority of SARS-CoV-2 infections to manifest atypical non-respiratory presentations, or even at times remain asymptomatic (*3*). Such minimally symptomatic individuals, never to be suspected of COVID-19, lack the opportunity to undergo SARS-CoV-2 nucleic acid amplification testing (NAT), and together with those false-negative for NAT, continue to carry the risk of becoming a source of transmission. COVID-19, being an unprecedentedly heterogenous pathology, constitute a spectrum of disease resembling an “iceberg”. Behind the most severe, devastating pneumonia patients lies the large majority that are only mildly symptomatic or even remain asymptomatic (*4*). Thus, NAT alone prone to overlooking the hidden burden, multi-tiered testing with the use of variable diagnostic modalities shall aid in exhaustive case detection along the whole spectrum.

The incidence as well as the origin of pauci-symptomatic or asymptomatic individuals forming a significant portion of the COVID-19 iceberg remain to be fully elucidated. In this study, 414 HCWs of a tertiary care hospital in Japan were tested for SARS-CoV-2 specific antibody seroconversion after facing an outbreak during the first wave of the pandemic in April-May 2020. The now-unveiled, overall perspective of the COVID-19 iceberg highlights the shocking underestimation of true disease burden, and holds an important lesson to be learned in minimizing nosocomial spreads and further enhancing preparedness against future pandemics.

## Materials and Methods

### Cohort and samples

A total 414 HCWs of St. Marianna University School of Medicine, Yokohama City Seibu Hospital, Kanagawa, Japan, who gave consent to participating in the study were recruited. Sera were obtained from the entire cohort within three consecutive days, from June 30^th^ to July 2^nd^ 2020, when approximately two months had passed since experiencing the nosocomial outbreak during April-May 2020. Amongst the individuals with a known date of COVID-19 diagnosis, the interval between the date of diagnosis and the date of serum sampling ranged from 6–10 weeks.

### Molecular testing

NAT for SARS-CoV-2 detection was performed using nasal swabs based on the RT-PCR protocol developed by the National Institute of Infectious Diseases, Japan. The method targets two sites of the nucleocapsid gene (*5*).

### Serological testing

Two chemiluminescent immunoassays, the Abbott SARS-CoV-2 IgG and SARS-CoV-2 IgG II Quant (Abbott, Illinois, USA), designed to detect serum IgG antibodies targeting the nucleocapsid and the spike proteins of SARS-CoV-2, respectively, were performed in accordance with the manufacturer’s instructions. A signal equal to or above a cutoff of 1.4 Index (S/C) and 50 AU/mL, respectively, was considered serologically positive. An orthogonal testing algorithm was adopted in order to idealize positive-predictivity and determine, with high specificity, the individuals who were truly sero-positive of SARS-CoV-2 specific antibodies (*3*). In this algorithm the individuals who initially tested positive for anti-nucleocapsid antibodies were tested with a second test targeting the SARS-CoV-2 spike antigen. Participants positive for both SARS-CoV-2 specific antibodies were finally confirmed of COVID-19 serological diagnosis by detecting neutralizing antibodies against SARS-CoV-2 using the Genscript SARS-CoV-2 sVNT (Genscript, Leiden, Netherlands), a competition ELISA-based surrogate virus neutralization assay. An inhibition rate (%inhibition) of 30% or above, which, according to the manufacturer’s instructions, is predictive of a half-maximal plaque reduction neutralization titer of 20 or higher, was selected as cutoff to determine positivity for neutralizing antibodies.

### COVID-19 case definition

Participants were defined as definitive COVID-19 patients, when either; (i) positive for NAT (“NAT-confirmed COVID-19”), or (ii) confirmed of positive serology by the orthogonal testing algorithm (“Serologically confirmed COVID-19”).

### Questionnaire for procedural exposure risk assessment

Participants completed a questionnaire which included demographic data, past medical history, occupational exposure to aerosol-generating procedures performed on confirmed COVID-19 patients, presence/absence of symptoms compatible with COVID-19, and state of NAT diagnosis. The procedural exposures of interest in this study were participation in (a) airway suctioning, (b) non-invasive ventilation (NIV), (c) bag mask ventilation, (d) nebulizer administration, (e) sputum induction, (f) oxygen supplementation as part of tracheostomy care, (g) endotracheal intubation/extubation, (h) tracheostomy, (i) bronchoscopy, and (j) cardiopulmonary resuscitation.

### Statistical analysis

The results of molecular or serological testing were described as frequencies and percentages among the participants screened. To assess the differences among demographic characteristics between NAT-confirmed and serologically confirmed COVID-19 patients, the following demographic variables were compared by t-tests (for the “Age” variable) or Fisher’s exact test (for the other variables; “Male sex”, “Pre-existing risk condition”, “Severity” and “Signs and symptoms”). Magnitude of serological response against the nucleocapsid and spike antigens, and the %inhibition surrogate virus neutralizability were compared by Mann-Whitney’s test according to symptom category; participants carrying respiratory and/or other systematic symptoms (“Symptomatic”), expressing no symptoms (“Asymptomatic”) and complaining of isolated smell impairments (“Hyposmia/anosmia only”). Spearman’s correlation coefficient was calculated for the various indices of serological response. For the procedural exposure risk assessment, risk ratio (RR), and risk difference (RD), per exposure were calculated as the ratio, or the absolute difference, between COVID-19 incidence among those exposed to the aerosol-generating procedures and the reference (“Not exposed”) group. The association between exposures to aerosol-generating procedures and COVID-19 incidence was tested by Fisher’s exact test. To evaluate the extent of harm attributable to each procedure regarding the actual increase of COVID-19 cases, the attributable fraction among the exposed (AFe) and the attributable number of events (AN) were calculated. AFe is the proportion of COVID-19 diagnoses in the exposed group that is attributable to the occupational exposure and was calculated per exposure as; AFe = (RR-1) / RR (*6*). AN is the absolute number of COVID-19 diagnoses attributable to the occupational exposure and was calculated per exposure as; AN = Afe × (number of COVID-19 diagnoses among the exposed). P-values less than 0.05 were considered statistically significant.

## Results

### Antibody seroconversion elucidates the true burden of the nosocomial outbreak underestimated by symptom-driven NAT screening

Of the 414 eligible and consented HCWs, 186 of 414 (44.9%) had underwent NAT screening for SARS-CoV-2 during the active emergence of the hospital cluster infection during April-May 2020. At the time, the approach towards screening of at-risk HCWs for COVID-19 was symptom-driven, and thus the participants who had never underwent NAT were those less prioritized due to either their lacking typical manifestations, or occupational exposures to aerosol-generating procedures performed on suspected/confirmed COVID-19 patients. 37 (19.9% of those tested by NAT and 8.9% of the entire HCW cohort) tested positive for SARS-CoV-2.

Approximately two months after the nosocomial outbreak had subsided, sera were collected from the participants and tested under the orthogonal testing algorithm (Figure 1). NAT and serological testing results are summarized in Table 1. Combining the NAT-confirmed and serologically confirmed diagnoses, the total number of COVID-19 cases and the overall prevalence rate summed to 64 and 15.5% (64/414), respectively.

**Table 1.** Summary of testing results

|  |  | *Serological testing* |  |  |
| --- | --- | --- | --- | --- |
|  |  | Positive | Negative | Total |
| *NAT* * | Positive |  |  | 37 |
|  | Negative | 23 | 126 | 149 |
|  | Not available | 4 | 224 | 228 |

\*Nucleic acid amplification test

**Figure 1.**
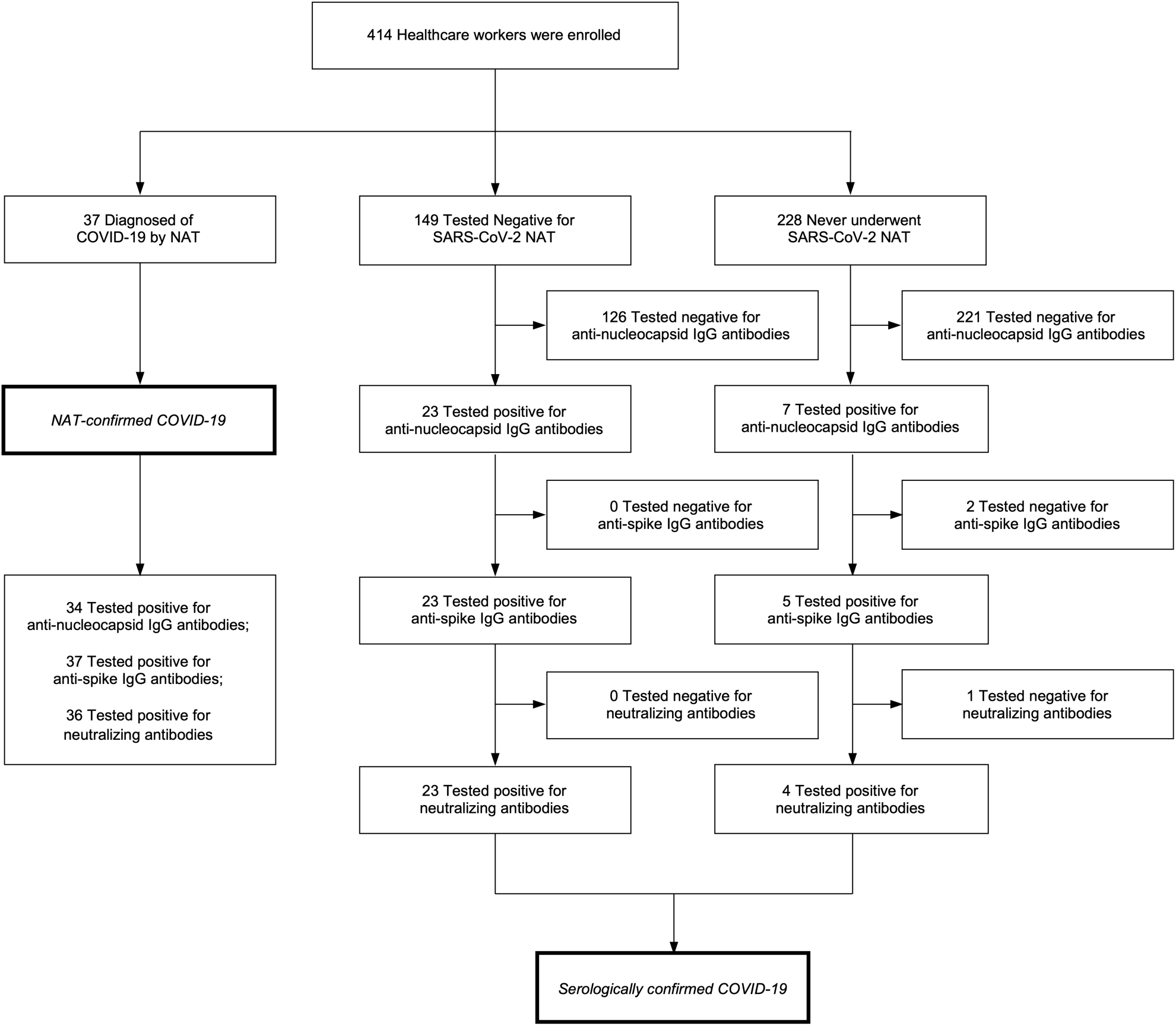
Enrollment, results of testing and algorithm for diagnosis. Of the 414 eligible and consented participants, 186 had underwent NAT testing for SARS-CoV-2. A total 37 of 186 tested healthcare workers were positive for NAT. The orthogonal testing algorithm led to the detection of 27 excess COVID-19 cases, diagnosed serologically. With NAT-and serology-confirmed cases combined, the total number of COVID-19 diagnoses summed to 64. NAT = Nucleic acid amplification test.

Symptom-driven NAT screening had overlooked 42.2% (27/64) of the definitive COVID-19 diagnoses. Of those serologically diagnosed, 23 of 27 (85.2%) had received negative NAT results, and 4 of 27 (14.8%) had been never suspected of COVID-19 and thus had not undergone NAT screening. After excluding those four individuals never having been tested by NAT, the sensitivity of NAT in COVID-19 case detection resulted to be as low as 61.7% (37/60).

### Clinical presentation, mode of diagnosis, and magnitude of serological response among the COVID-19 HCW cohort

Demographic data from the COVID-19 cases within the HCW cohort of the present study is demonstrated in Table 2. The mean age was 35 (±12) years and 11 of 64 (17.2%) were male. Only 4 of 64 (6.3%) had known high-risk comorbidities (hypertension and/or diabetes) and 4.7% (3 of 64) reported chronic steroid use. Regarding the severity of disease, the majority of symptomatic COVID-19 cases were mild to moderate illnesses and only 1 of 64 (1.6%) required O_2_ supplementation, with no case fatality reported. Typical respiratory symptoms were present in 31 of 64 (48.4%) of the COVID-19 cases and others presented with isolated hyposmia/anosmia (6 of 64, 9.4%) or less specific systemic symptoms; headache, abdominal symptoms and/or malaise (8 of 64, 12.5%). Notably, all six cases presenting with isolated hyposmia/anosmia were confirmed by NAT (6 of 6, 100%). To the contrary, asymptomatic cases (19 of 64, 29.7%) were mainly confirmed by serological testing (17 of 19, 89.5%).

**Table 2.**
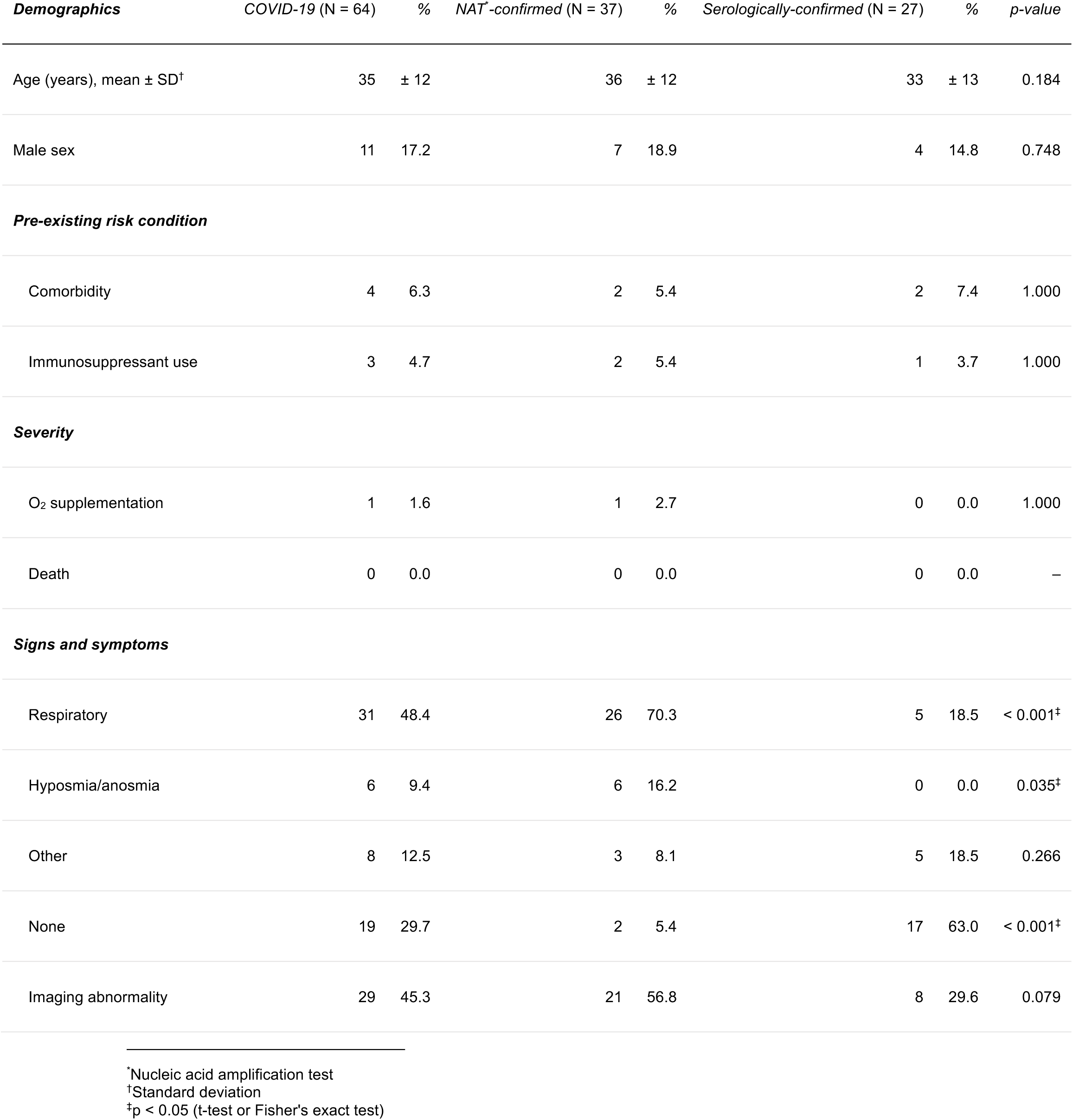
Participant demographics

Cross quantitative comparison of the elicited immune responses (Figure 2, panel A) showed that the magnitude of immune response targeting the two major nucleocapsid and spike antigens showed significant correlation within an individual (Spearman’s r = 0.666, p < 0.0001). Further, compared with the levels of anti-nucleocapsid antibody titer (Spearman’s r = 0.560, p < 0.0001), a stronger correlation was observed between anti-spike antibody titers and surrogate virus neutralizability (Spearman’s r = 0.857, p < 0.0001) (Figure 2, panel B). Interestingly, compared with the other symptom categories, participants presenting with isolated hyposmia/anosmia elicited anti-nucleocapsid and anti-spike antibody responses of significantly lower magnitude, constituting an immunologically distinct subpopulation (Figure 3, panel A). Similarly, competition ELISA-based surrogate virus neutralization assay showed a trend towards lower neutralizability of the “hyposmia/anosmia only” subpopulation, though not reaching statistical significance (Figure 3, panel B).

**Figure 2.**
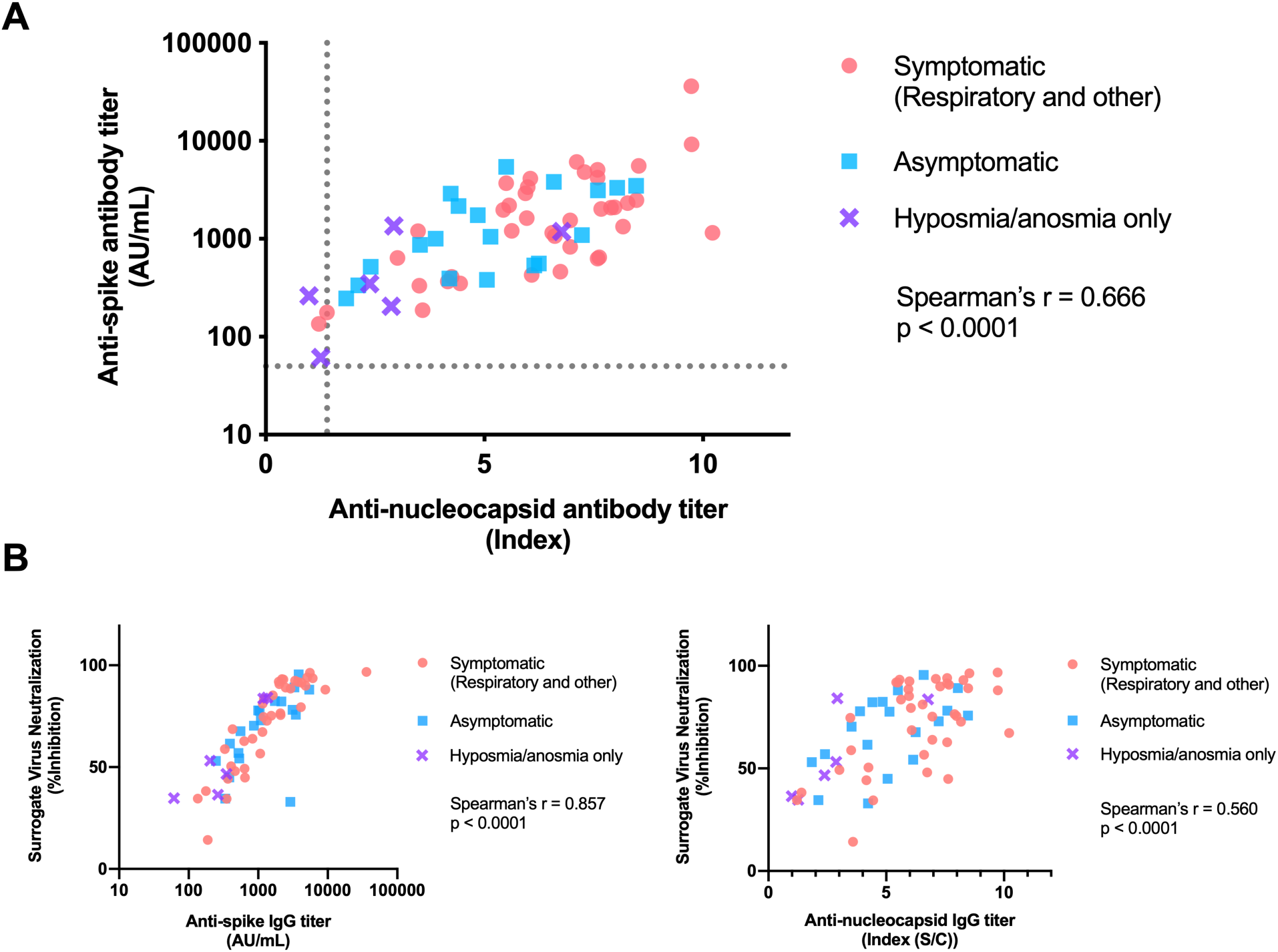
Quantitative assessment of serological responses and their mutual relationships. A) Magnitude of serological response against the two major SARS-CoV-2 antigens. Dotted lines indicate cutoff values. B) In comparison to the anti-nucleocapsid IgG titer, the level of SARS-CoV-2 neutralizability, assessed by the surrogate virus neutralization assay, was correlated to a stronger extent with the anti-spike IgG titer.

**Figure 3.**
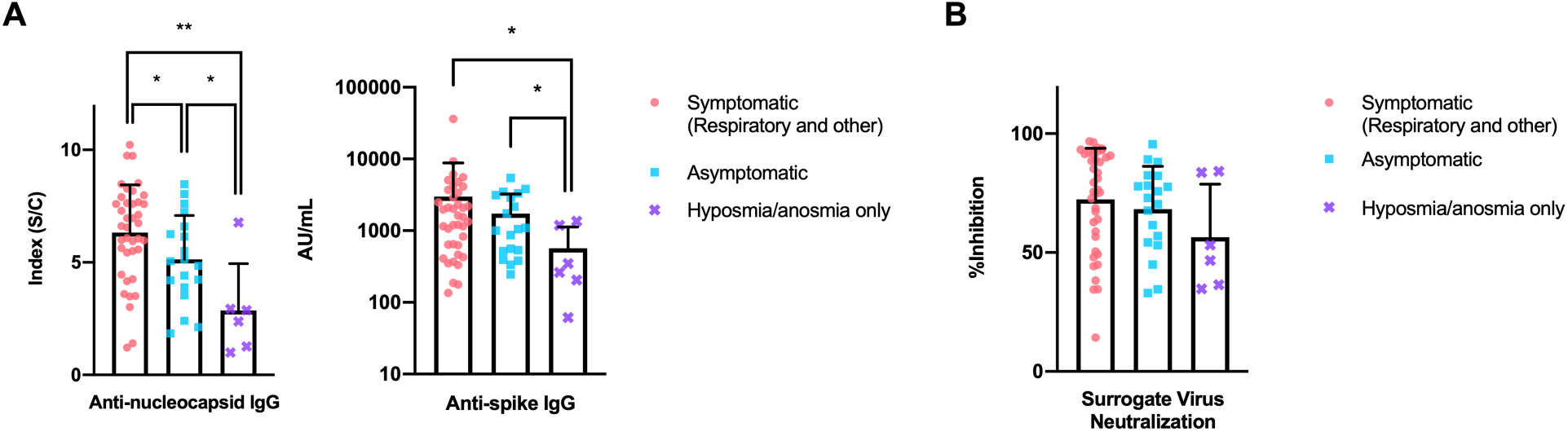
Serological status of SARS-CoV-2 affected healthcare workers by symptom category. A) Healthcare workers with COVID-19 diagnosis that had manifested isolated hyposmia/anosmia were characterized by diminished serological responses against the two major SARS-CoV-2 antigens. B) The similar trend towards lower SARS-CoV-2 neutralizability of sera obtained from the 6 participants with isolated hyposmia/anosmia did not reach statistical significance. ^*^p < 0.05, ^**^p < 0.01 Mann-Whitney’s test.

### Defining procedural exposure-related risks

Of the 414 eligible participants, 212 (51.2%) reported to have had participated in aerosol-generating procedures and thus had experienced SARS-CoV-2 exposures (Table 3). Amongst the variable types of aerosol-generating procedures, NIV (RR 3.10, p = 0.008) conveyed the highest risk of SARS-CoV-2 transmission to the exposed HCWs, followed by airway suctioning (RR 1.67, p = 0.040). Although sputum induction and cardiopulmonary resuscitation also seemed to convey substantial transmission risks to the exposed, the present study was underpowered to observe statistical significance in the risk increase related to these exposures.

**Table 3.**
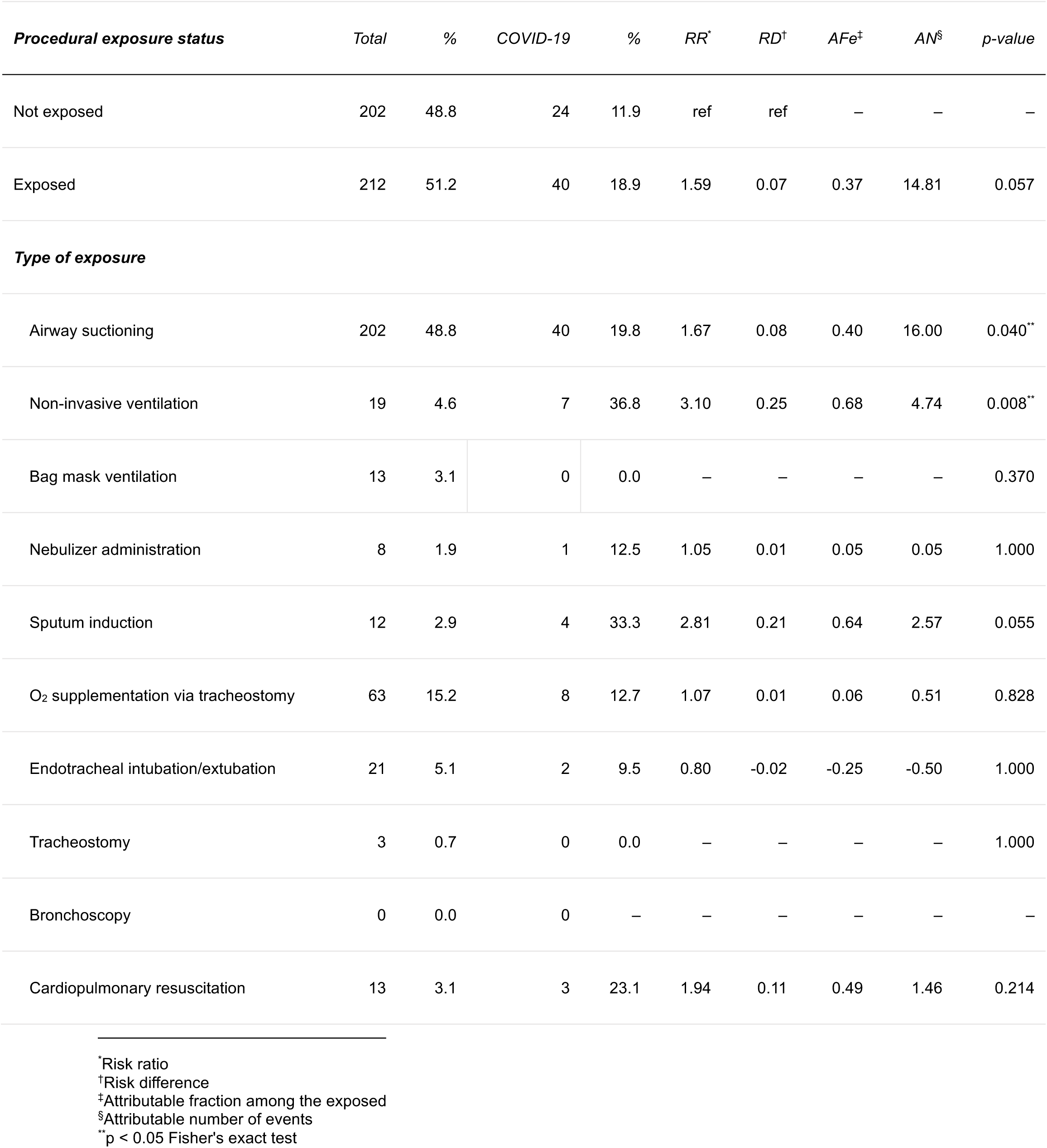
Risk of SARS-CoV-2 transmissibility during exposure to aerosol-generating procedures

Although the procedural risk inherent to airway suctioning seemed substantially lower compared with NIV, airway suctioning, being a commonly performed aerosol-generating procedure, was the exposure to which the highest number of excess COVID-19 cases were attributed (Table 3).

## Discussions

The composite approach of combining NAT and serology-based diagnoses exhaustively detected definitive COVID-19 cases in the Japanese HCW cohort experiencing a nosocomial outbreak during April-May 2020. A surprising 42.2% of overlooked COVID-19 diagnoses had occurred when case detection had relied solely on NAT, leading to undetected transmission, and triggering the outbreak. Keeping this iceberg phenomenon in mind, appropriate allocation of testing resources is needed in order to effectively detect contagious individuals, clarify the true burden of COVID-19, and eradicate transmission in nosocomial settings.

NAT-based case detection in Japan had been counted on as a promising strategy, capable of thoroughly tracking SARS-CoV-2 transmissions, and identifying and sizing cluster infections (*1*). It was not until June 2020, when the first national seroprevalence survey was performed, that the Japanese realized their 3–8 fold underestimation of the actual spread of the disease within the society (*3*). With the aim of enhancing case detection for effective quarantine, especially among the pre-symptomatic or asymptomatic affected individuals, testing recommendations since then have shifted from a symptom-driven approach towards a rather universal approach. Against expectations, however, having been the sole first-tier diagnostic against this emerging infection, it is now increasingly recognized that NAT-based SARS-CoV-2 pathogen detection faces serious limitations. COVID-19 illness being primarily a lower respiratory tract infection, the probability of pathogen detection from upper respiratory tract specimens decrease rapidly and nearly halves within approximately two weeks from onset (*7*). Previous reports have suggested that a substantial fraction, as high as up to 54%, of COVID-19 patients may present with undetectable viral loads and show false-negative RT-PCR results (*8–10*). Our observation recapitulates such findings, by demonstrating the sensitivity of NAT to have remained as low as 61.7%. Missed diagnoses having occurred not only in the pauci-symptomatic and the asymptomatic populations but also in acutely ill cases of high suspicion, indefinite molecular testing results already have left behind a significant burden of those in need of a diagnosis. A well-defined diagnostic complementary to NAT is still in serious need.

Since the host immune response lags behind viral invasion, the ability of antibody tests to detect an acute infection in its early phase is usually limited and considered inferior to NAT. However, in the case of COVID-19, NAT performance itself remains suboptimal and thus serological testing may well aid in early-phase case detection (*11,12*). While the present study targeted pre-exposed HCWs and was designed so as to establish delayed COVID-19 diagnoses, accumulating evidence further supports the clinical usefulness of serological testing in acute care and diagnosis. COVID-19 pneumonia with repeatedly false-negative NAT results, is not an uncommon clinical scenario, where serological testing, having an extended detectable window, may work complementarily and establish the diagnosis in the early phase of illness (*11*). Used in combination with NAT, serological testing has proven to enhance case detection and help control the spread of SARS-CoV-2 when applied to carefully targeted, high-risk populations, such as in-hospital outbreaks resembling the HCW cohort of the present study (*12*). By the use of chemiluminescence immunoassays as applied in the present study, exerted sensitivities may rise nearly as high as 40% and 80% by day 7 and day 14 of illnesses, respectively (*13,14*). Therefore, the performance of well-designed platforms may potentially serve as comparable alternatives to NAT in the very acute phase (day 4– 7) and may even outperform NAT in the later phases (beyond day 10). In addition, COVID-19 related long-lasting sequelae, such as anosmia or the multisystem inflammatory syndrome in children are widely accepted suitable indications for serological testing (*15,16*). Limitations in capacity for molecular testing, still an ongoing issue in resource-deprived settings, is another factor which makes serological diagnostics an attractive alternative for acute diagnosis purposes.

In addition, exhaustive case detection has here enabled precision of risk estimates innate to aerosol-generating procedures. Our observations are in support of the prevailing concerns on the risks that aerosol-generating NIV may create for HCWs and provide implications regarding the origin of nosocomial spreads (*17*). Notably, while the procedural risk inherent to airway suctioning seemed substantially lower compared with NIV, airway suctioning, being a commonly performed aerosol-generating procedure, was the exposure to which the highest number of excess COVID-19 cases were attributed. The above findings warn frontline HCWs about the harms of undervaluing risks related to any specific procedural exposure and stress once again the importance of being equipped with appropriate protectives when confronting novel pathogens.

Cross-comparison of the participants’ serological responses has highlighted the heterogeneity in width and magnitude of humoral immune responses among the affected. The observed higher detection rate of isolated hyposmia/anosmia patients by NAT, and the uniquely suppressed humoral immune response of the subpopulation may be reflecting confined viral replication and subsequent localized host immune reactions to the nasal airway. However, to draw conclusions on the relationships between viral tropism and serological responses of the host, data laying emphasis on individuals presenting with isolated hyposmia/anosmia are still lacking. Therefore, it remains a future consideration to refine pretest probabilities and to individualize diagnostic approaches based on case presentation (*18,19*).

In conclusion, by way of analyzing serological status against SARS-CoV-2, we unveiled the missed diagnoses within HCWs from a tertiary care hospital in Japan, which had experienced a COVID-19 outbreak during the first peak of the pandemic. Our observations here emphasize the efficiency of well-designed serological diagnostics in the detection of COVID-19 cases and SARS-COV-2 transmissions, and indicate that the true spread within the hospital was almost twice as extensive than previously estimated using a symptom-based NAT surveillance. Multi-tiered diagnostics are key to tracing the exact shape of the COVID-19 iceberg and without the consideration of the hidden but significant portion of the iceberg beneath the surface, we face the risks of under-estimating COVID-19 disease prevalence, over-estimating death rates, and misinterpreting exposure-specific risks.

## Data Availability

The data that support the findings of this study are available from the corresponding author upon reasonable request.

## Acknowledgments

This research was supported by Japan Agency for Medical Research and Development (AMED) under Grant Number JP20wm0125003 (YK), JP20he1122001(YK), JP20nk0101627(YK) and JP20jk0110021 (YN). The authors receive financial support from the Special Reserves Fund for COVID-19 (Osaka City University) and the COVID-19 Private Fund (Shinya Yamanaka Laboratory, Center for iPS Cell Research and Application, Kyoto University). YN is a recipient of the BIKEN Taniguchi Scholarship. Minako Hosokawa, Hiroko Tanaka, Tomoyo Tominaga, Harumi Domyo from the St. Marianna University School of Medicine, Yokohama-city Seibu Hospital, supported the questionnaire distribution and sample/data collection. Reagents for serological testing were provided from Abbott Japan LLC, Japan.

## Ethical Statement

Analyses were conducted in accordance with the ethical standards noted in the 1964 Declaration of Helsinki and its later amendments. The research was approved by the institutional ethics committee (#2020-003). Consent for participation and publication was obtained from every participant.

